# Associations Between Perfusion Index During Out-Of-Hospital Cardiopulmonary Resuscitation, Cardiopulmonary Resuscitation Quality and Return Of Spontaneous Circulation

**DOI:** 10.1101/2025.08.01.25332657

**Authors:** Stefano Malinverni, Simon Kalisz, Marta Frigerio, Carolina Cogozzo, Gaia Pedicini, Garance Clignez, Marie-Astrid de Villenfagne, Dina Farhat

## Abstract

**BACKGROUND:** The association between perfusion index (PI) and out-of-hospital cardiac arrest survival outcomes is unknown. The American Heart Association suggested the need to improve the monitoring of cardiopulmonary resuscitation and chest compression quality. We hypothesized that higher event-average perfusion index values are associated with higher probability of return of spontaneous circulation, better survival and neurological outcomes.

**METHODS:** In this prospective cohort study of index out-of-hospital cardiac arrests attended from January 2022 through October 2024, we analyzed the association of the event-average perfusion index value with sustained return of spontaneous circulation. Secondary exposures included survival to hospital admission, favorable neurological outcome (Cerebral Performance Category ≤3 or no change from baseline) and associations between epoch-average PI and CPR quality metrics.

**RESULTS:** We included 98 index out-of-hospital cardiac arrests (mean [Standard Deviation]) age 62.1 years [17.6], 32% female, 9.2% shockable rhythm. Median event-average perfusion index was 0.29 ([Q1, Q3], 0.11, 0.92). Events with sustained return of spontaneous circulation had a higher event-level average perfusion index (0.92 [0.44, 1.73] mmol/L versus 0.19 [0.09, 0.55]; *P*<0.001). The receiver operating characteristic analyses of PI, as a predictor of sustained return of spontaneous circulation, identified an area under the curve of 0.77 [95% CI, 0.68– 0.86]) with an optimal cut point at 0.61 (sensitivity 0.67, specificity 0.81). After adjusting for confounders, PI was associated with higher relative risk of sustained ROSC (adjusted odds ratio, 2.6 [95% CI, 1.3–5.1]; *P*<0.001). Event-average PI was also associated with higher relative risk of survival to hospital admission (adjusted odds ratio, 2.1 [95% CI, 1.2–3.7]; *P*=0.001) while it was not associated with higher relative risk of neurological intact survival (adjusted odds ratio, 4.0 [95% CI, 0.8– 19.4]; *P*=0.085). Higher epoch-average PI was independently associated with higher chest compression fraction (*P*=0.003).

**CONCLUSIONS:** Higher event-average PI was associated with higher probability of sustained return of spontaneous circulation. Event-average PI was associated with higher adjusted probability of return of spontaneous circulation, but not with neurological intact survival. Epoch-average PI was positively correlated with chest compression fraction.

## Introduction

Approximately 33% of patients will achieve a return to spontaneous circulation (ROSC) and 8% will be discharged home alive after out-of-hospital cardiac arrest (OHCA).[1]

Given the low survival rates, the high estimated cost of unsuccessful resuscitation[2] and the limitations of simple pre-hospital termination of resuscitation (TOR) rules[3], there is a need to develop tools to monitor and titrate the resuscitation effort according to physiologic metrics, to assist the decision to end resuscitation and objectively assess futility.

End-tidal carbon dioxide (ETCO2) provides a non-invasive estimate of cardiac output and organ perfusion during cardiac arrest [4,5] and its use is recommended to monitor the quality of cardio-pulmonary resuscitation (CPR)[6]. Low ETCO2 values during resuscitation reflect the low cardiac output generated by chest compressions [7]. ETCO2 may serve as a non-invasive hemodynamic indicator, albeit with complexities of interpretation[8].

During CPR, ETCO_2_ values depend both on the blood flow generated by chest compressions, on minute alveolar ventilation and on metabolic activity of patient tissues[9,10]. Studies on ROSC detection and patient outcome rely on the comparison of measured ETCO2 levels[11]. However, chest compression quality— measured by rate and compression depth—and ventilation parameters such as rate and tidal volume, influence ETCO2 during CPR with opposing effects [12,13] thus complicating its interpretation. Other limitations are the need of endotracheal tube placement to accurately measure ETCO2 and its discordant reduction relative to coronary perfusion pressure following the administration of adrenaline[14]. Given the conflicting results on the ability of ETCO2 to predict outcomes on an individual-patient basis and the limitations in its interpretation induced by minute ventilation during CPR further investigations and the development of new tools are needed in this area[15].

The perfusion index (PI) is the ratio of pulsatile, on non-pulsatile light absorbance of the photoplethysmography signal. The determinants of PI are complex, reflecting the interaction between peripheral and central haemodynamic factors, such as vascular tone and stroke volume[16]. PI correlates with stroke volume, mean arterial pressure and cardiac index with increases in each parameter being associated with corresponding increases in PI[17–19]. Conversely, PI is negatively correlated with hypovolemia and increased vascular tone. Thus, PI represents a FDA-approved measure of peripheral perfusion that can be rapidly, continuously and non-invasively obtained via a pulse oximeter, without the need for an advanced airway that could be used to monitor CPR.

Perfusion index has been associated during post-ROSC with 30-day survival [20] with higher values associated with increased survival rates. However, its utility as a predictor of ROSC during CPR remains poorly investigated.

Establishing PI targets during resuscitation could be a practical means of broadly monitoring resuscitation quality and possibly improving outcomes.

To address the knowledge gap we prospectively designed an observational cohort study to evaluate the association between PI during CPR and: (1) ROSC (2) survival outcomes, and (3) CPR quality markers.

We hypothesized that higher event-average of PI during OHCA CPR would be associated with a higher likelihood of ROSC, a greater incidence of neurological intact survival, and superior CPR quality metrics.

## Methods

### Study design and setting

This study is a registered prospective observation cohort of an a priori defined sample of 92 consecutive non-traumatic OHCAs (NCT05383885). It was conducted between January 2022 and October 2024 in patients attended by the mobile intensive care unit (MICU) of Centre Hospitalier Saint-Pierre in Belgium.

The institutional review board of the CHU Saint-Pierre hospital approved the study (CE/21-11-01).

The data that support the findings of this study are available from the corresponding author on reasonable request.

### Subject population

Patients victims of OHCA older than 11 years requiring chest compressions for at least 4 minutes were included. The age threshold was determined as expected body weight and size of 12 years old subjects allows the utilization of adult sensors for perfusion index measurement.

Subjects were excluded if they: (1) were pregnant, (2) were prisoners (3) achieved return of circulation through venoarterial extracorporeal membrane oxygen support (4) did not have at least 4 minutes of perfusion index data recorded (two 2-minutes epochs).

### Data collection

We collected standardized cardiopulmonary arrest data elements, CPR quality mechanics data (compression depth and rate), electrocardiogram monitor data and quantitative capnography for CPR events. As soon as feasible, without interfering with the quality of ongoing CPR, patients were monitored using a Rad-97 Pulse CO-Oximeter by Masimo applied at the fingertip, preferably the middle finger, of the hand opposite the intravenous cannula. The finger was shielded from ambient light using a self-adhesive bandage and stabilized alongside an adjacent finger to minimize motion artifacts.

ETCO2 measurements were performed using a cap-ONE mainstream probe integrated within a Corpuls3 monitor.

Chest compression frequence and depth as well as ECG curves were retrieved from the Corpuls3 monitor.

Perfusion index sampled at a frequency of 0.5 Hz and averaged on 2-minutes epochs as well as ETCO2 values, also averaged on 2-minutes epochs were deidentified and subsequently reviewed and analyzed. Investigators reviewed each epoch for critical features including: (1) the start of CPR, (2) interruptions in CPR, (3) periods of nonsustained ROSC, and (5) sustained ROSC.

A minimum of 4 minutes and up to the entire duration of CPR was collected, comprising perfusion index and ETCO2 values during resuscitation (up to 22 epochs of 120 seconds each). No epochs were excluded from the analysis. Absence of PI values was interpreted as a signal in itself, and a PI value of 0 was assigned in such cases.

PI values were not hidden from the treating physician but no decision, during the arrest and post-arrest care of the participants, was taken on treatment based on the perfusion index values. All patients received standard ACLS as recommended by the American Heart Association guidelines and the decision to stop resuscitation procedures were made in accordance with recommendations for Resuscitation Termination Rules (TOR) in cardiac arrest[21].

Demographic data, the cause of arrest and cardiac arrest timeline were based on the Belgian Cardiac Arrest Registry (B-CAR), a national registry that follows Utstein recommendations approved by each participating site ethical committee and of the Free University of Brussels acting as the academical coordinating center. Baseline modified Rankin Scale, information on sustained ROSC and outcome Cerebral Performance Category were extracted from the B-CAR and assessed by clinicians independent of to the study.

All monitoring modalities were recorded simultaneously until ROSC or death as determined by the treating physician.

Periods of intermittent ROSC were excluded from the analysis. The event average of each physiological variable was the average of all evaluable epochs averages for a given case. If a patient experienced a period of unsustained ROSC, the same patient was analysed for the two reanimation efforts; the first one ending with an unsustained and the second one ending with either sustained ROSC or death.

### Exposures and endpoints

We prospectively defined the primary aim as investigating the association of the event average of PI and sustained ROSC. The study was registered before enrolling the first patient (NCT05383885).

The primary exposure was the event average of PI. The primary outcome was sustained ROSC, defined as a spontaneous circulation for more than 20 minutes since the interruption of chest compressions[22].

In the absence of previous studies we calculated the sample size a priori hypothesizing a sensitivity and specificity of 90% for PI to predict ROSC, an expected prevalence of ROSC of 40%, a precision of 10% and a type I error rate of 0.05. Based on these assumptions and on a possible dropout of 5% we calculated a final sample size of at least 92 resuscitations attempts.

Secondary outcomes included admission alive to the hospital and neurological intact survival defined as a cerebral performance score below 3 or unchanged from baseline. Intra-arrest CPR mechanics and quality variables included: (1) chest compression depth (mm), (2) chest compression rate (min–1), (3) chest compression fraction (proportion of cardiopulmonary arrest event during which chest compressions are provided), (4) ETCO2 (mm Hg).

### Statistical analysis

Patient and event characteristics were summarized according to occurrence of sustained ROSC. Summaries reported were counts and percentages for categorical variables, mean±SD for normally distributed continuous variables, or median (quartile 1, quartile 3) for non-normally distributed continuous variables.

Associations of summarized variables with PI were assessed with Fisher exact test for nominal variables and with the Wilcoxon rank-sum test for continuous variables that were not normally distributed. Receiver operating characteristic curves (ROC) were used to examine the performance characteristics of prognostic tests and the area under the ROC curve was used to assess performance. The optimal cutoff point for PI to predict ROSC was determined from the ROC curve by the Youden index without consideration of covariables. Using the identified cut-off we sought to evaluate PI as a dichotomized rather than continuous exposure. CPR mechanics as well as outcomes were summarized by PI group. We used directed acyclic graphs (ie, causal diagrams) to pre-identify potentially important confounders that were visualized using the DAGitty online tool (Figure S1). Multivariable logistic regression models examined associations between PI and sustained ROSC adjusting for the previously identified confounders: age, initial rhythm of VF/VT, mean chest compression depth, chest compression fraction and low-flow duration.

Several exploratory analyses were conducted. First we conducted the ROC analyses to study PI predicting value in terms of admission alive to the hospital and neurological intact survival. Second, multivariable logistic regression models examined the association between PI and hospital admission alive or neurological intact survival adjusting for the previously identified confounders. Third, epoch-level CPR quality and physiological data were summarized by PI group. Associations of summarized variables with PI groups were assessed with the Wilcoxon rank-sum test for continuous variables. A multivariable regression using epoch-level data was created to investigate the relationship of PI with: (1) CCF, (2) chest compression depth, (3) first observed rhythm and (4) no-flow. Fourth, to provide a useful bedside marker of probability of ROSC, the trend in PI over the last 10 minutes of CPR was depicted in patients with and without ROSC. The mean values were compared between patients with and without ROSC through mixed-effects linear regression modelling with a gamma distribution of PI with constant values replacement for null values and a random subject effect to give address correlation between 2 different minutes of CPR for the same subject.

A two-sided *p* value of less than 0.05 was used to establish statistical significance. No adjustment for multiple comparisons was performed. Analysis was performed using Stata 16.1 (StataCorp, College Station, TX, USA).

## Results

Among the 248 OHCA attended by our MICU during the study period 138 were included in the study. In 131 cases the device was not used. Due to a technical malfunction of the study device 15 patients were not included and 10 were excluded according to the specified exclusion criteria (Figure 1). In total, 30 patients attained sustained ROSC; survival to hospital discharge was 9.2% and survival with favorable neurological outcome was 5.1%.

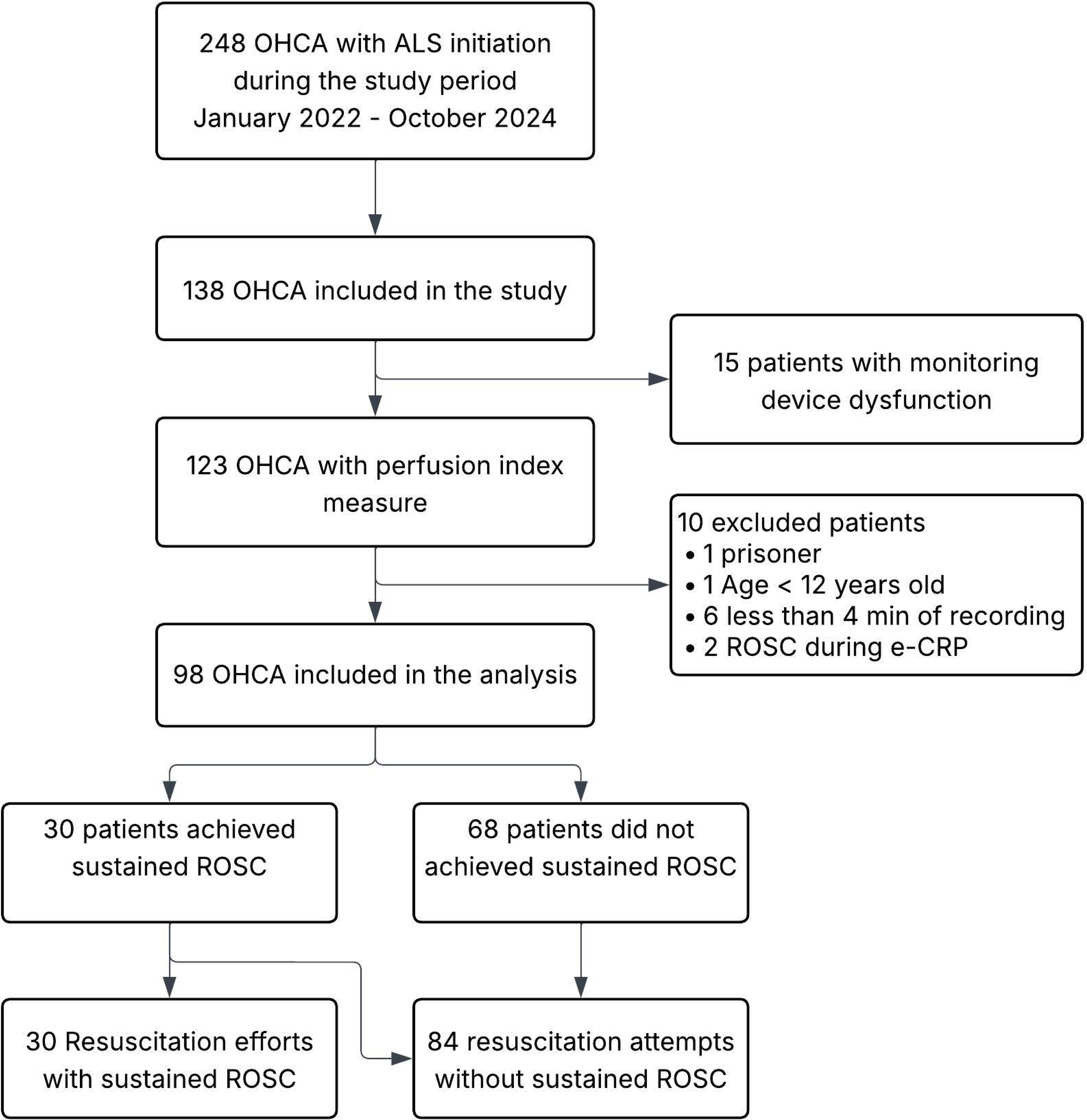

Overall patients characteristics as well as summarized by PI group are presented in Table 1. Median (quartile 1, quartile 3) event-average PI was 0.29 (0.11, 0.92). In terms of preexisting conditions before CPR, the two groups had similar characteristics. Overall event characteristics as well as summarized according to ROSC are presented in Table 2. CPR events with sustained ROSC had a higher event-level average PI (0.92 [0.44, 1.73] mmol/L versus 0.19 [0.09, 0.55]; *P*<0.001; Figure 2).

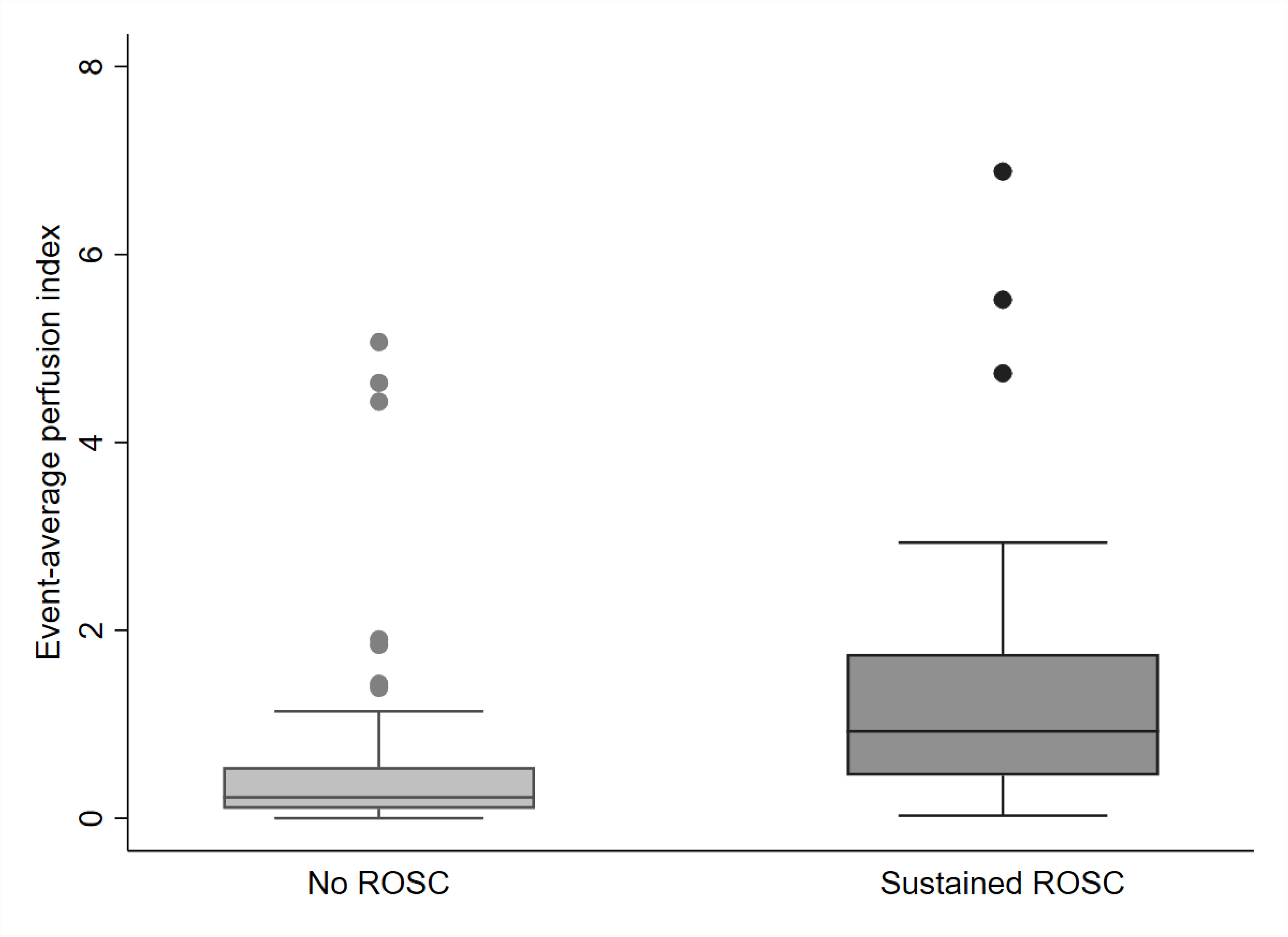

Figure 3 illustrates the receiver operating characteristic curve the relationship between PI and ROSC. The receiver operating characteristic analyses of PI, as a predictor for sustained ROSC, identified an area under the curve of 0.77 [95% CI, 0.68–0.86]) with an optimal cut point at 0.614 (sensitivity 0.67, specificity 0.81).

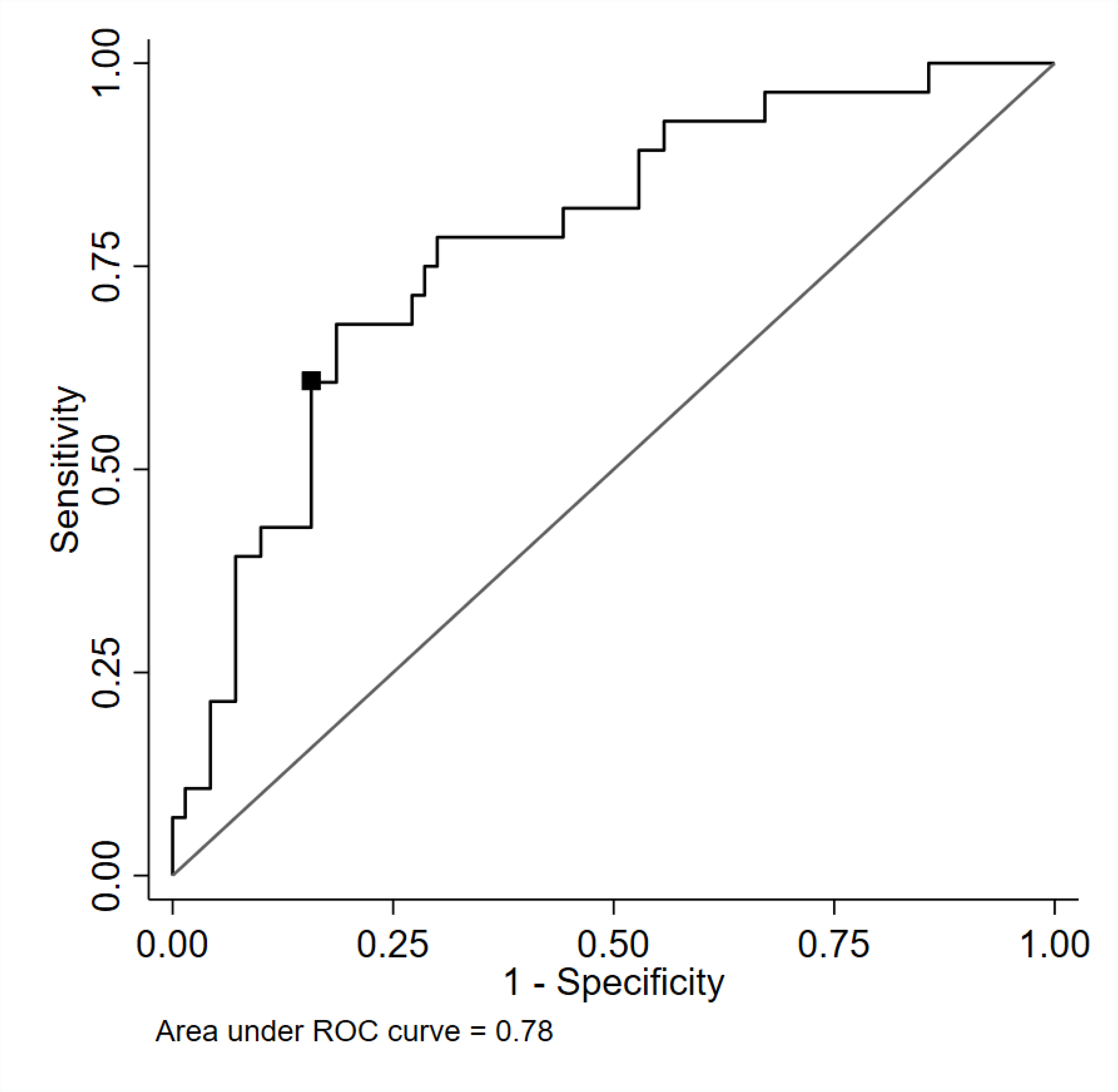

Out of 114 resuscitation attempts, 37 (32.5%) had an event-level average ≥0.614. More patients had ROSC when event-level average PI was ≥0.614 compared with lower PI values (24/37 [64.9%] versus 15/77 [19.5%]; *P*<0.001). Multivariable outcome adjusted analyses of event-level average PI as a predictor of sustained ROSC and other secondary outcomes are presented in Table 3. After adjusting for confounders, a PI remained associated with higher relative risk of sustained ROSC (adjusted odds ratio, 2.6 [95% CI, 1.3–5.1]; *P*=0.005).

### Exploratory Analyses

The receiver operating characteristic analysis of PI, as a predictor for admission alive to the hospital and neurological intact survival are presented in Figures S2 (survival to hospital admission) and S3 (survival with good neurological outcome). The ROC analysis of PI as a predictor of hospital admission alive identified an area under the curve of 0.69 [95% CI, 0.59–0.79]) with an optimal cut point at 0.614 (sensitivity 0.56, specificity 0.83). The ROC analysis of PI as a predictor of neurological intact survival identified an area under the curve of 0.74 [95% CI, 0.53–0.94]) with an optimal cut point at 0.211 (sensitivity 1, specificity 0.44).

After adjusting for confounders, event-average PI was associated with higher relative risk of survival to hospital admission (adjusted odds ratio, 2.1 [95% CI, 1.2–3.7]; *P*=0.01) while it was not associated with higher relative risk of neurological intact survival (adjusted odds ratio, 4.0 [95% CI, 0.8–19.4]; *P*=0.085) (Table 3).

CRP characteristics and metrics by PI group, at an epoch-level, are presented in Table 4. Epochs with average PI≥0.614 had a higher chest compression rate (114 [110, 120] compressions/minute versus 110 [108, 115] compressions/minute, *P*<0.001). A multivariate analysis correcting for quality of CPR metrics, no flow and the first presenting rhythm, found that PI was positively correlated with CCF (*P*=0.003) but not with chest compression depth or frequency.

**Table 4:**
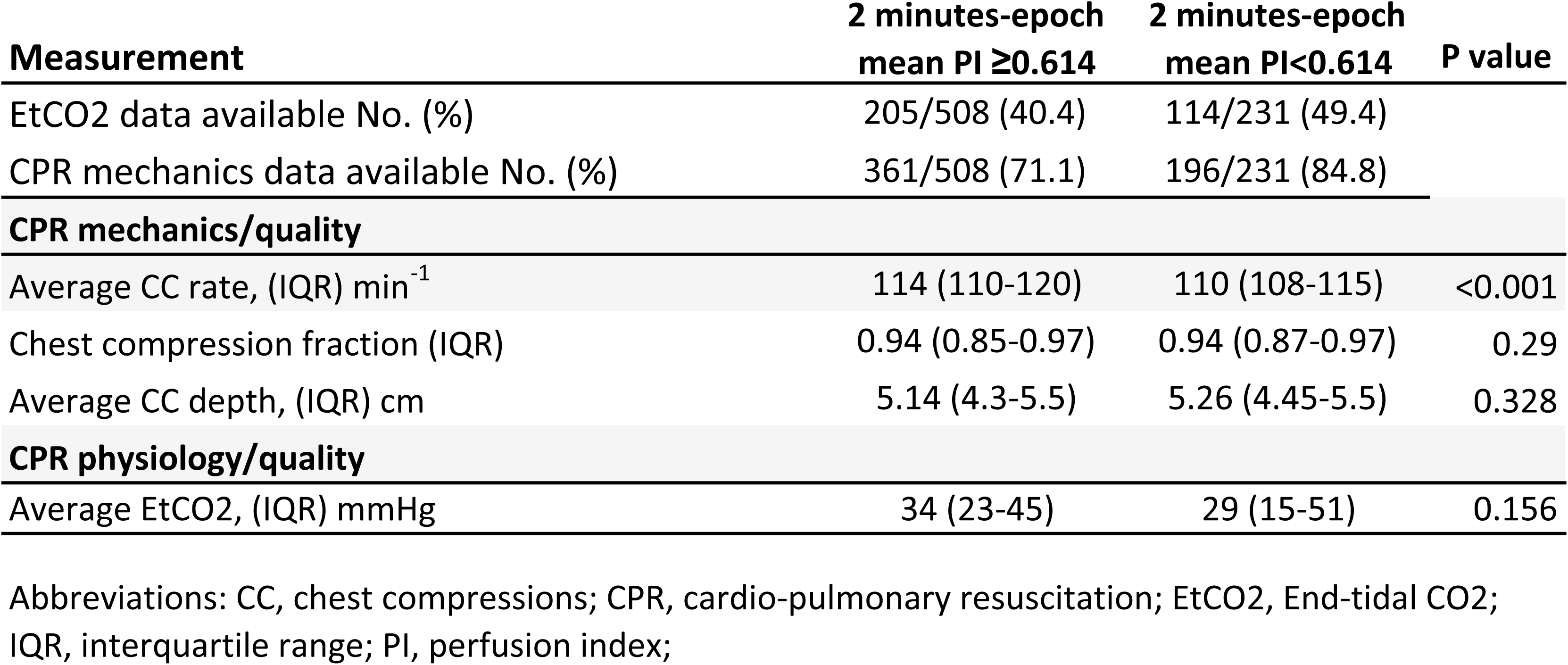
CPR Quality and physiology.

The trend in PI during the last 20 minutes of CPR is graphically represented in Figure S5. There was a significant difference in the epoch-level mean PI (P<0.001) but not in the slope of PI change over time (effect size estimate, -0.03 [95% CI, - 0.07–0.01]; P=0.012) among patients achieving sustained ROSC versus those who did not.

## Discussion

In this single-center cohort study of adult patients with out-of-hospital cardiopulmonary arrest who had a non-invasive measure of PI during CPR, higher PI during CPR was associated with a higher incidence and adjusted relative risk of ROSC and hospital admission alive. An event-average PI≥0.614 mm Hg during CPR was identified as a possible threshold as it was associated with a higher incidence and adjusted relative risk of ROSC. To our knowledge, this is the first clinical study providing evidence to support monitoring PI during CPR or provide a PI target for CPR.

Our study provides the first evidence to identify an association of PI with ROSC and survival to hospital admission. It demonstrates a significant finding suggesting that PI might be associated with CPR metrics during CPR by showing the positive correlation between PI and higher chest compression fraction during CPR, a primary driver of blood flow during cardiac arrest.

The International Liaison Committee on Resuscitation, supports the idea that rescuers should monitor and titrate the resuscitation effort according to CPR metrics and a patient’s physiological response. Moreover, given the lack of conclusive evidence on the subject, it recommends further research to identify new tools and targets for optimizing CPR[23]. Adjusting the CPR technique (e.g., depth and position of chest compressions) and vasopressor administration according to either invasive arterial blood pressure[24] or ETCO2[25] is associated with better cardiopulmonary arrest outcomes in large animal models. Although prospective clinical observational studies have established ETCO2 targets, and diastolic pressure targets during CPR both measurements are associated with limitations. Invasive blood pressure is intrinsically difficult to measure during OHCA. ETCO2 is associated with multiple limitations such as the need of an invasive airway to precisely measure reliable values[26] and the technical difficulties associated with continuous capnometry (obstruction from tracheal tube secretions and disconnection). ETCO2 measurements were available, in a pediatric arrest study conducted in the ICU, in less than two thirds of patients already monitored and intubated before arrest[27], suggesting that ETCO2 monitoring is less available in OHCA. Moreover ETCO2 is inversely associated with minute ventilation [8].

Consequently, when interpreting ETCO2 during CPR, minute alveolar ventilation acts as a significant confounding factor[10]. Finally, chest compression induced oscillations in the ETCO2 signal may limit ETCO2 interpretation and reliability[28] as these are associated with ample variations in the recorded ETCO2, making the highest value of expired CO2 during oscillations, and not ETCO2, the best estimate of alveolar CO2[28].

In an effort to identify an additional clinically useful metric for out-of-hospital care providers, we investigated the prognostic value of a ubiquitous, inexpensive, simple, easily applied, and non-invasive parameter. Animal models had demonstrated that both the area under the curve and the amplitude of the pulse oximetry plethysmographic waveform correlate with chest compression depth, coronary perfusion pressure, and the PI[29] while human studies found an association between pulse oximetry plethysmography and ROSC [30]. Although a multi-center study demonstrated a relationship between the plethysmographic signal and ROSC, it implied manipulations of the plethysmographic signal data with post-hoc computations that reduced its clinical application and relevance. PI shares similarities with the area under the curve of the plethysmographic waveform, as both are influenced by the intensity of the plethysmographic signal. PI measurement does not require complex ex-post computations and can be readily displayed on standard monitors, offering the potential to provide prognostic information and eventually guide CPR in real time at the bedside.

PI is associated with stroke volume[31], mean arterial pressure[18], vascular tone[32] and volume status[17]. Each of these parameters is an important determinant of cardiac output during CPR. PI could therefore non-invasively monitor important hemodynamic factors and represent not only a prognostic factor but also a potential target on which titrating CPR intervention. PI presents challenges in interpretation and is subject to limitations, including its correlation with extremity temperature[33] and decreased accuracy in the presence of motion artifacts[34], which are common during CPR. In order to minimize motion artifacts, we used self-adhesive single use sensors, light shields and recommended a syndactyly between the finger with the sensor and an adjacent finger to improve the noise-to-signal ratio.

A previous prospective, observational study of a mixed population of out-of-hospital and in-hospital cardiac arrests found no association between PI and ROSC[35]. This cohort studied 1-min epoch at 5, 10, 15, 20 and 25 minutes after intubation and compared PI values for each epoch which might have introduced a survivorship bias in the analysis. Conversely, a multicentric study demonstrated that the peripheral perfusion index averaged over 30 minutes post-ROSC could discriminate 30-day survival with good neurological outcomes, providing further supporting evidence for its use in cardiac arrest beyond the CPR period[36].

Our study, together with previous evidence demonstrating an association between the PI and multiple determinants of ROSC, suggests that PI may serve as a complementary tool to ETCO2 and CPR quality metrics—such as CCF and compression depth during CPR. PI provides non-invasive monitoring of the interplay between stroke volume and vascular tone during chest compressions [16], and may function as a prognostic indicator of the likelihood of achieving ROSC.

This study has limitations. First, the observational study design precludes our ability to assign causation between higher PI levels and improved outcomes. Yet, our findings provide support to suggest a direct mechanistic relationship between higher PI values and outcomes through an improved chest compression fraction. Second, these data were collected as part of a single center initiative, which was conducted in a tertiary-care MICU within a two-tier response system for OHCA. These characteristics, must be considered before generalizing our findings across diverse care environments. Third, none of the included patients had a measurement of PI in place at the start of CPR, and as such, whether our findings can be extrapolated to the early phases of CPR remains an unanswered question. Finally, due to known intrinsic difficulties in the study of physiological variables in OHCA, we report data from 41.2% of patients who were eligible for inclusion, raising concerns regarding selection bias.

## Conclusions

In this single center study, an higher event-average PI during CPR was associated with higher risk of sustained ROSC in out-of-hospital cardiopulmonary arrest, providing support for further research that advise monitoring PI during CPR.

## Data Availability

Data will be available from the corresponding author upon reasonable request

